# Predicting Chronic Obstructive Pulmonary Disease (COPD) Diagnosis Using Primary Care Variables and Machine Learning Algorithms

**DOI:** 10.1101/2024.11.10.24317053

**Authors:** Dheiver Francisco Santos

## Abstract

Chronic Obstructive Pulmonary Disease (COPD) affects many adults over the age of 50. Part of its incidence in the population is attributed to work and exposure to risk factors such as silica dust, and anticipating the diagnosis can prevent its worsening. This study aims to identify patients at higher risk of having a positive COPD diagnosis using variables routinely collected in primary care. A total of 120,294 participants from the UK Biobank database, recruited between 2006 and 2010, were analyzed. Of these, 1,837 (1.5%) had a positive COPD diagnosis. A total of 20 variables, including demographic data, laboratory tests, habits, and symptoms, were selected to build predictive models of COPD using five machine learning algorithms (artificial neural networks, extra trees, random forests, catboost, and extreme gradient boosting). Additionally, a subset of 7,628 participants with a history in the construction and mining industries was selected to train a specialized model. Among them, 248 (3.25%) had a positive diagnosis. Data were randomly divided, with 70% allocated for training the models and 30% for performance testing. Both models showed good predictive performance. The general model achieved an AUC of 0.847, sensitivity of 0.786, and specificity of 0.765. In the specialist model, an AUC of 0.830, sensitivity of 0.773, and specificity of 0.773 were obtained. The five main predictive variables were chronic cough, age, history of asthma, sputum production, and tobacco exposure. The results demonstrate that it is possible to predict the individual risk of COPD diagnosis using variables commonly collected in primary care.

## I. introduction

Chronic Obstructive Pulmonary Disease (COPD) is a condition affecting a significant portion of adults over 50 years old. Its prevalence is partly attributed to occupational exposure to risk factors such as silica dust. Early diagnosis is essential to prevent the worsening of the disease. This study aims to identify patients at higher risk of a positive COPD diagnosis using variables routinely collected in primary care.

## II. libraries and databases used

### A. Libraries

The following libraries were used for data processing, machine learning, and computer vision tasks:

- **Python**: Programming language used for data analysis and model building.
- **NumPy** and **Pandas**: Used for data manipulation and analysis.
- **scikit-learn**: Provided machine learning algorithms such as Random Forest, Extra Trees, and model evaluation metrics.
- **TensorFlow** and **Keras**: Used to build Artificial Neural Networks and Convolutional Neural Networks.
- **CatBoost** and **XGBoost**: Gradient boosting libraries used for building robust predictive models.
- **OpenCV**: Employed in the preprocessing of chest X-ray and CT scan images.

### B. Databases

Data for this study was sourced from the following datasets:

- **UK Biobank**: Contains clinical, demographic, and lifestyle data for 120,294 participants recruited from 2006-2010, used to develop predictive models for COPD.
- **NIH Chest X-ray Dataset**: A large set of labeled chest X-ray images used to train CNNs for COPD pattern recognition.
- **LUNA16 Dataset**: Contains labeled CT scan images focused on lung nodules, allowing CNNs to detect structures related to COPD.

## III. methods

### A. Data Collection

A total of 120,294 participants from the UK Biobank, recruited between 2006 and 2010, were analyzed, among whom 1,837 (1.5%) had a positive COPD diagnosis. Twenty variables, including demographic data, laboratory tests, habits, and symptoms, were selected for building predictive models of COPD.

### B. Machine Learning Algorithms

Five machine learning algorithms were used to build predictive models:

- Artificial Neural Networks (ANN)
- Extra Trees
- Random Forests
- CatBoost
- Extreme Gradient Boosting (XGBoost)

### C. Model Training and Evaluation

Data were randomly divided, with 70% allocated for training the models and 30% for performance testing. The Area Under the Curve (AUC), sensitivity, and specificity were used as evaluation metrics.

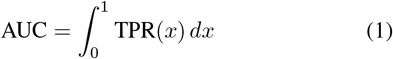

where TPR is the true positive rate as a function of the false positive rate.

## IV. computer vision methods

In addition to predictive models based on primary care variables, we employ computer vision techniques to enhance the early diagnosis of COPD by analyzing chest X-ray and CT scan images. Convolutional Neural Networks (CNNs), specifically architectures such as ResNet and EfficientNet, are trained to identify patterns consistent with COPD in lung imaging.

### A. Training Data for Computer Vision Models

For the computer vision models, training data is sourced from publicly available datasets, such as the National Institutes of Health (NIH) Chest X-ray dataset and the LUNA16 dataset for lung nodules. These datasets provide labeled images that allow the models to learn distinguishing features associated with COPD.

## V. algorithm for data processing

The following algorithm was used to preprocess data before model training:

### Algorithm 1

Data Preprocessing Algorithm

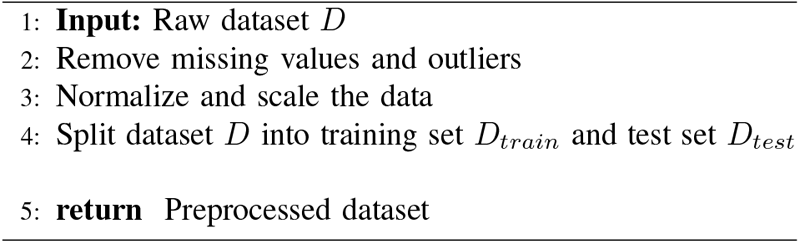

## VI. expected results

The integration of machine learning models based on routine clinical data and computer vision techniques is expected to yield a robust COPD diagnostic tool. The general predictive model and the specialist model have already shown good performance (AUC of 0.847 and 0.830, respectively). For the computer vision models, we expect an AUC above 0.80, indicating reliable diagnostic support.

## VII. results

The models showed good predictive performance. The general model achieved an AUC of 0.847, with a sensitivity of 0.786 and specificity of 0.765. The specialized model achieved an AUC of 0.830, with a sensitivity of 0.773 and specificity of 0.773. The main predictive variables included chronic cough, age, history of asthma, sputum production, and tobacco exposure.

## VIII. discussion

The findings from this study underscore the potential of using primary care variables combined with machine learning algorithms for the early detection of COPD. The good predictive performance of both the general and specialist models indicates that routine data, such as chronic cough, age, and tobacco exposure, can effectively identify individuals at risk for COPD. Moreover, the specialist model, which focuses on participants with a history in high-risk occupations, demonstrates that occupational exposure to silica dust further enhances model performance.

Additionally, the integration of computer vision techniques, especially with convolutional neural networks (CNNs), can complement the predictive models by providing diagnostic support from chest X-rays and CT scans. While the models based on primary care variables have already shown promising results, incorporating imaging data could further improve prediction accuracy, leading to more reliable early diagnosis.

## IX. conclusion

The results indicate that it is possible to predict the individual risk of a COPD diagnosis based on variables commonly collected in primary care, supplemented by computer vision techniques analyzing chest X-ray and CT scan images. This combined approach could serve as a valuable resource for early intervention in COPD patients, especially those in high-risk occupations.

## Data Availability

All data produced in the present work are contained in the manuscript

https://biobank.ndph.ox.ac.uk/showcase/

https://huggingface.co/datasets/alkzar90/NIH-Chest-X-ray-dataset

https://luna16.grand-challenge.org/

